# Algorithmic design choices as determinants of health equity: a causal simulation study of healthcare resource allocation

**DOI:** 10.64898/2026.10.30.26366732

**Authors:** Chayce Reed, Monica E. Espinoza

**Affiliations:** Department of Biomedical Data Science, Geisel School of Medicine, Dartmouth College, Hanover, NH, USA

**Keywords:** algorithmic fairness, care management, health equity, resource allocation, causal inference, All of Us Research Program

## Abstract

**Background:** Care management programs increasingly rely on prediction algorithms to decide which patients receive limited healthcare resources. Because these algorithms are typically trained to predict future healthcare utilization or cost, they may systematically underidentify patients whose care-seeking behavior is suppressed by structural barriers.

**Objective:** To evaluate how three modifiable design choices (selection threshold, feature set, and training objective) affect allocation equity in care management targeting.

**Methods:** We constructed an analytical cohort of 201,216 adults from linked electronic health record and participant survey data in the NIH All of Us Research Program (2017-2019). Three prediction models (logistic regression, random forest, and XGBoost) were trained and evaluated under simulation conditions that varied one design choice at a time while holding the others fixed. Equity was assessed using false negative rate (FNR), equal opportunity difference (EOD), and allocation rate conditional on need (an access-independent benchmark based on self-reported general health status). We also applied Oaxaca-Blinder decomposition to identify which predictors contributed most to the Black-White allocation gap.

**Results:** Prior utilization understated self-reported health burden among disadvantaged groups: 33% of Black and 32% of Hispanic participants reported fair or poor health, yet only 22% and 19% were classified as high utilizers. White participants showed the opposite pattern: 25% were classified as high utilizers, while only 19% reported fair or poor general health status. Under baseline design choices, all three models produced similar disparities, with uninsured participants showing the largest gap (EOD: -0.148). Oaxaca-Blinder decomposition identified prior outpatient visits as the dominant contributor to the Black-White gap in predicted allocation, with higher average emergency department utilization among Black participants partially offsetting the effect. Varying the selection threshold had little effect; disparities persisted across all thresholds. Feature set variation produced mixed and group-specific effects; removal of prior outpatient visits achieved near-zero Black-White EOD (0.001, AUC: 0.77), consistent with the Oaxaca-Blinder decomposition finding. Training objective variation produced the most substantial and consistent equity effects, with the health status objective achieving near-zero Black-White EOD (0.008, AUC: 0.69) and the survey-enriched health status condition reversing the direction of disparity for most groups.

**Conclusions:** Algorithmic equity in care management targeting is largely a consequence of modifiable design choices, with training objective emerging as the most impactful of the three design choices evaluated. The causal simulation framework and access-independent equity benchmark developed here provide a replicable approach for evaluating equity as a design consideration during algorithm development rather than a post-deployment correction.

## 1. Introduction

Health systems increasingly rely on algorithmic tools to identify patients for care management programs [1–3]. These programs offer intensive services, such as care coordination, medication management, and social support, to patients expected to have high future healthcare costs and needs [4, 5]. Central to their design is a risk scoring algorithm that is trained on administrative claims or electronic health record data and uses prior healthcare utilization, costs, diagnoses, and demographic characteristics to predict which patients will have the highest utilization or cost in a future period [1, 6].

A growing body of evidence has documented that algorithmic tools used in healthcare can produce racially and socioeconomically disparate outcomes, systematically underidentifying high-need patients from disadvantaged groups [2, 7–9] In a landmark analysis, Obermeyer and colleagues demonstrated that a widely deployed commercial algorithm used to identify patients for care management programs exhibited racial bias, assigning lower risk scores to Black patients than to White patients with equivalent health burden [2]. The authors concluded that the disparity stemmed from the algorithm’s reliance on prior healthcare costs as a proxy for health need. Utilization-based measures reflect not only health burden but also differential access to care, and therefore systematically underestimate need among populations whose utilization is suppressed by structural barriers [7, 10, 11]. This finding has since been replicated and extended across clinical settings and algorithm types [8, 12, 13], establishing algorithmic bias in healthcare resource allocation as a pressing and pervasive problem.

Despite growing awareness of algorithmic bias in healthcare, existing research has focused primarily on documenting its existence rather than systematically evaluating the design choices that produce it [14]. Furthermore, prior work has largely evaluated algorithmic equity using measures derived from the same utilization data used to train the models, limiting the ability to detect disparities that arise precisely because structural barriers make observed utilization an inadequate proxy for health need [2, 15]. Therefore, a systematic evaluation of how modifiable algorithmic design choices independently shape equity outcomes is needed, and should be assessed against an accessindependent measure of health need.

This study evaluates how three modifiable design choices in care management targeting affect allocation equity: the selection threshold, the feature set, and the training objective. Using data from the NIH All of Us Research Program, and grounding the analysis in a structural causal model of the allocation system, we treat each design choice as an intervention on the system’s causal graph, enabling systematic evaluation of each choice’s independent effect on equity.

We make four contributions. First, we develop and apply a causal simulation framework for auditing algorithmic design choices, providing a replicable approach to evaluating design effects on equity in observational healthcare data. Second, we leverage the linkage of electronic health record data with self-reported health status from the All of Us participant survey to evaluate allocation equity against an access-independent measure of health need, capturing disparities that utilization-based metrics cannot detect in populations whose healthcare-seeking behavior is suppressed by structural barriers. Third, we apply Oaxaca-Blinder decomposition to identify which predictor characteristics contribute to the observed allocation disparity, and in what proportion. Fourth, we quantify the direction and magnitude of equity effects associated with each design choice across demographic and socioeconomic subgroups, characterizing which interventions shift allocation equity most and at what cost to predictive accuracy.

## 2. Methods

### 2.1. Study design

We conducted a decision simulation study to examine how algorithmic design choices in care management targeting affect equity across demographic and socioeconomic groups. Using a structural causal model (SCM) framework, we treated each design choice as an intervention on the allocation system’s causal graph, enabling direct evaluation of three modifiable algorithmic decisions: selection threshold, feature set, and training objective.

### 2.2. Data source

We used data from the NIH All of Us Research Program, a longitudinal precision medicine initiative enrolling a diverse cohort of U.S. adults, with a deliberate commitment to including historically underrepresented populations in biomedical research. All of Us links electronic health record (EHR) data from participating health systems with participant-reported survey data collected at enrollment, allowing clinical utilization patterns to be paired with self-reported health status and socioeconomic characteristics. Data was accessed through the Researcher Workbench, a cloud-based analysis environment providing registered users access to participant-level data under an approved data use agreement.

### 2.3. Study cohort

We identified a cohort of adults aged 18 or older at baseline (January 1, 2017) who had at least one inpatient, emergency department, or outpatient visit during a two-year baseline period (2017-2018) and completed the All of Us enrollment survey during 2017-2019. Healthcare utilization was measured as the count of distinct visit dates across inpatient, emergency department, and outpatient encounter types. Participants were required to have non-missing data on the primary utilization outcome and the general health status equity benchmark. Missing values for body mass index and income were retained in the analytical cohort using binary missing indicators as additional predictors, rather than excluding participants with incomplete data on these variables. All other predictor variables in the survey-enriched feature set were required to be non-missing, resulting in a common analytical cohort used consistently across all simulation conditions to ensure comparability.

### 2.4. Outcome variable

The primary outcome was high healthcare utilization in the follow-up year (2019), defined as a binary indicator for membership in the top quartile of total visit counts (inpatient, emergency department, and outpatient visits combined), consistent with fixed-proportion targeting approaches used in deployed care management programs in which a predetermined share of patients is identified for resource allocation based on predicted utilization risk [1, 2].

### 2.5. Equity benchmark

We evaluated allocation equity using self-reported general health status as an access-independent measure of health need, where participants reporting fair or poor general health status were classified as having high health need. Unlike prior utilization, which is subject to access barriers that may suppress healthcare-seeking behavior among disadvantaged populations, self-reported general health status reflects participants’ own assessment of their health burden independent of their ability to access care. This benchmark enables evaluation of whether equally sick participants from different demographic and socioeconomic groups received resources at equal rates.

### 2.6. Feature sets

We evaluated four feature sets: standard, outpatient restricted, full utilization restricted, and survey-enriched. The standard feature set included demographic characteristics (age, sex), prior healthcare utilization (inpatient, emergency department, and outpatient visit counts during 2017-2018), chronic condition indicators (diabetes, hypertension, heart disease, chronic obstructive pulmonary disease, depression, obesity, cancer), chronic condition count, body mass index, and a missing indicator for body mass index. The outpatient restricted feature set excluded prior outpatient visits while retaining inpatient and emergency department visit counts. The full utilization restricted feature set excluded all prior utilization variables, retaining only demographic and clinical characteristics. The survey-enriched feature set extended the standard feature set with socioeconomic and social determinants of health variables derived from All of Us survey data, including self-reported household income, insurance type, housing instability, smoking history, alcohol use, and functional limitation status.

### 2.7. Models

We trained three prediction models representing commonly used approaches in care management targeting: logistic regression, random forest, and gradient boosted trees (XGBoost) [2, 16–18]. Logistic regression served as the primary interpretable baseline given its transparency and ease of clinical implementation relative to more complex model classes [18]. Random forest and XGBoost were included to evaluate the robustness of findings to model class. Random forest was fit with 500 trees. XGBoost was fit with 100 boosting rounds, a maximum tree depth of 6, and a learning rate of 0.1. All remaining hyperparameters were set to package defaults. Hyperparameters were held fixed across all simulation conditions so that observed differences reflect design choice effects rather than differences in model tuning. A fixed selection threshold was applied to each model’s predicted probability output to flag the top 20% of participants by predicted risk, reflecting a typical care management program capacity constraint [1, 2]. The analytical cohort was randomly partitioned into a 70% training set and 30% test set using stratified sampling on the high utilizer outcome. All performance and equity metrics were computed on the held-out test set.

### 2.8. Simulation framework

We evaluated three modifiable algorithmic design choices as interventions on the allocation system’s structural causal model (Figure 1).

**Figure 1:**
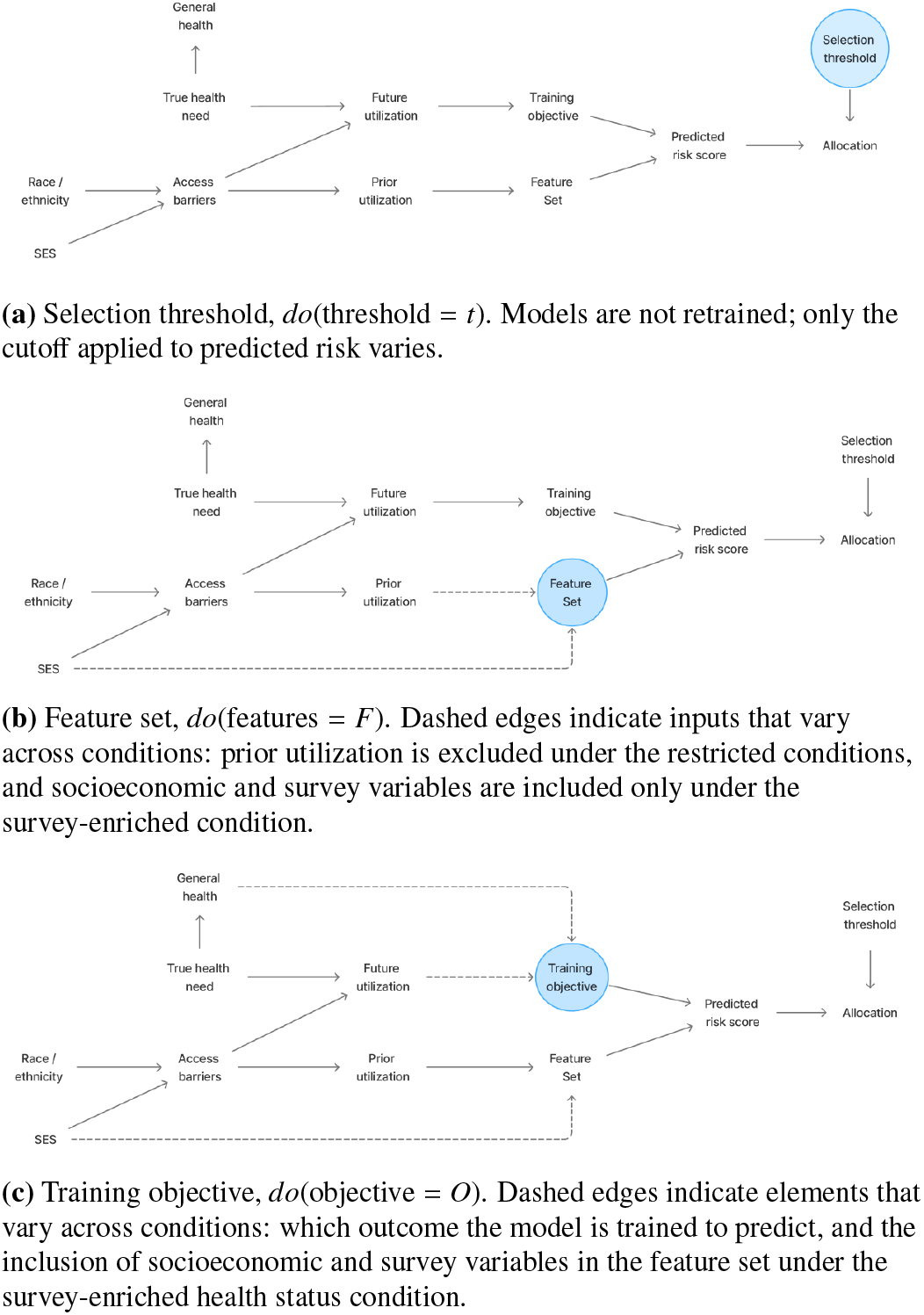
Each design choice as an intervention on the allocation system: (A) selection threshold, (B) feature set, and (C) training objective. Shaded nodes indicate the intervened variable in each condition.

Simulation 1 examined the effect of selection threshold by applying three thresholds (10%, 20%, 30%) to model predictions without retraining. This operationalizes the policy decision of how many participants to flag as *do*(threshold = *t*) (Figure 1a). The 20% threshold condition serves as the reference.

Simulation 2 examined the effect of feature set by retraining all three models under standard, outpatient restricted utilization, full utilization restricted, and survey-enriched feature set conditions, operationalized as *do*(features = *F*) (Figure 1b). The standard feature set condition serves as the reference.

Simulation 3 examined the effect of training objective by retraining all three models under four conditions operationalized as *do*(objective = *O*) (Figure 1c). The standard utilization condition trained models to predict top-quartile total healthcare utilization using the standard predictor set. The outpatientrestricted utilization condition trained models to predict topquartile inpatient and emergency department utilization using the standard predictor set. The standard health status condition trained models to predict fair or poor self-reported general health status using the standard predictor set. The surveyenriched health status condition trained models to predict fair or poor self-reported general health status using the surveyenriched predictor set. The standard utilization condition serves as the reference.

### 2.9. Equity metrics

We evaluated allocation equity using two complementary metrics computed separately for each subgroup variable (race/ethnicity, insurance type).

The primary equity metric was the false negative rate (FNR), defined as the proportion of actual high utilizers not flagged by the model, computed separately for each subgroup. A higher FNR for a given group indicates that the model systematically fails to identify high utilizers from that group, resulting in disproportionate exclusion from care management resources. To quantify that disparity, we computed the equal opportunity difference (EOD) for each subgroup, defined as the differences in true positive rates between each subgroup and its reference group [19–22].

The secondary equity metric was the allocation rate conditional on need, defined as the proportion of participants with fair or poor self-reported general health status flagged by the model, computed separately for each subgroup. This metric represents a novel contribution of this study, enabled by the linkage of EHR utilization data with All of Us participant survey data. Unlike FNR, which uses observed utilization as the definition of need, the allocation rate conditional on need uses self-reported health status as an access-independent benchmark, capturing disparities that arise when structural barriers suppress healthcare utilization for disadvantaged populations relative to their true health burden. To quantify disparity on this metric, we computed the allocation rate gap for each subgroup, defined as the differences in allocation rate conditional on need between each subgroup and its reference group.

For both metrics, reference groups were White for race/ethnicity and private insurance for insurance type. A negative EOD indicates higher FNR for the subgroup relative to the reference group. A negative allocation rate gap indicates lower allocation among fair or poor health participants relative to the reference group.

### 2.10. Disparity decomposition

To identify which predictor characteristics contribute to racial disparity in predicted allocation, we applied Oaxaca-Blinder decomposition [23] to the predicted log-odds of allocation from the logistic regression model, using the Black-White comparison as the primary illustrative case. The three-fold decomposition partitions the mean difference in predicted log-odds between two groups into an endowments component (reflecting differences in observed characteristics between groups), a coefficients component (reflecting differential weighting of those characteristics by the model), and an interaction component (capturing the joint effect of both). Because all participants were scored by the same fitted model and race was not among its predictors, the coefficients and interaction components were negligible, and the results are interpreted through the endowments component. Decomposition was conducted with White participants as the reference group and bootstrap standard errors estimated from 1000 replications.

### 2.11. Statistical analysis

All analyses were conducted in R (version 4.6.1) using the following packages: tidyverse (version 2.0.0) for data manipulation, caret (version 7.0.1) for stratified train/test partitioning, ranger (version 0.18.0) for random forest estimation, xg-boost (version 3.2.1.1) for gradient boosted trees, pROC (version 1.19.0.1) for AUC computation, oaxaca (version 0.1.5) for Oaxaca-Blinder decomposition, gtsummary (version 2.5.1) for cohort characteristic tables, and ggplot2 (version 4.0.3) for figures.

## 3. Results

### 3.1. Study cohort

The analytical cohort comprised 201,216 adults. Participants had a mean age of 50.3 years (SD 16.2) and were 62% female (Table 1). The cohort was racially and ethnically diverse, with 51% identifying as White, 21% as Black, 18% as Hispanic, 7.2% as Other or Multiple races, and 2.9% as Asian. By insurance type, 32% were privately insured, 20% enrolled in Medicaid, 18% in Medicare, 2.5% in VA coverage, and 0.7% uninsured; insurance type was unknown for 27%. Income distribution was skewed toward lower income levels, with 37% reporting annual household income below $35,000, 25% between $35,000 and $100,000, and 19% above $100,000; income was not reported for 19%. Among all participants, 23% met high utilizer criteria (top quartile of total visit count during 2019) and 25% reported fair or poor general health status. High utilizers were more likely to be older, have higher chronic disease burden, and report fair or poor general health status than standard utilizers. Body mass index was missing for 13% of participants, who were retained in the analytical cohort using a binary missing indicator.

**Table 1.**
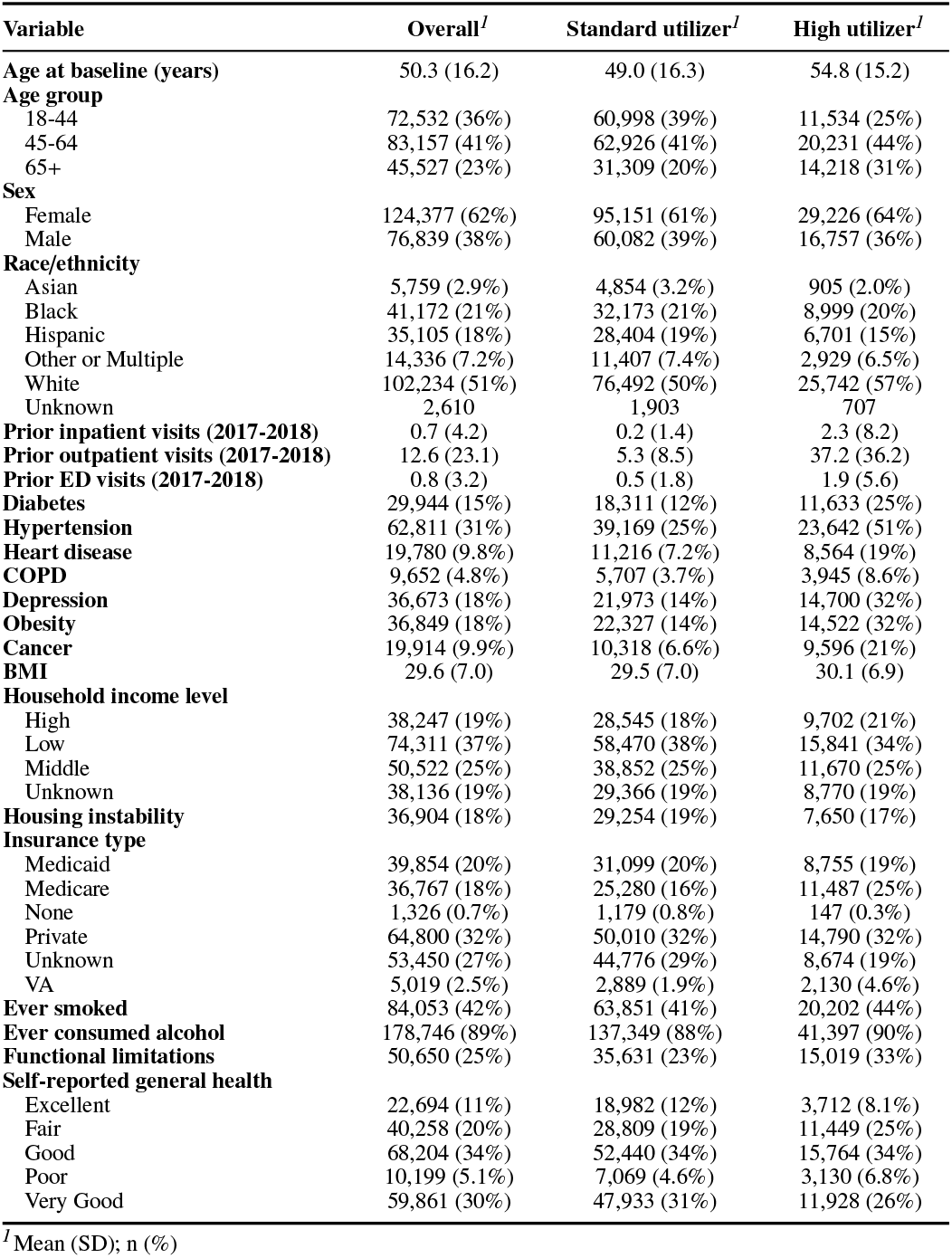
Cohort characteristics by high utilizer status.

| **Variable** | **Overall**<sup>*1*</sup> | **Standard utilizer**<sup>*1*</sup> | **High utilizer**<sup>*1*</sup> |
|---|---|---|---|
| **Age at baseline (years)** | 50.3 (16.2) | 49.0 (16.3) | 54.8 (15.2) |
| **Age group** |  |  |  |
| 18-44 | 72,532 (36%) | 60,998 (39%) | 11,534 (25%) |
| 45-64 | 83,157 (41%) | 62,926 (41%) | 20,231 (44%) |
| 65+ | 45,527 (23%) | 31,309 (20%) | 14,218 (31%) |
| **Sex** |  |  |  |
| Female | 124,377 (62%) | 95,151 (61%) | 29,226 (64%) |
| Male | 76,839 (38%) | 60,082 (39%) | 16,757 (36%) |
| **Race/ethnicity** |  |  |  |
| Asian | 5,759 (2.9%) | 4,854 (3.2%) | 905 (2.0%) |
| Black | 41,172 (21%) | 32,173 (21%) | 8,999 (20%) |
| Hispanic | 35,105 (18%) | 28,404 (19%) | 6,701 (15%) |
| Other or Multiple | 14,336 (7.2%) | 11,407 (7.4%) | 2,929 (6.5%) |
| White | 102,234 (51%) | 76,492 (50%) | 25,742 (57%) |
| Unknown | 2,610 | 1,903 | 707 |
| **Prior inpatient visits (2017-2018)** | 0.7 (4.2) | 0.2 (1.4) | 2.3 (8.2) |
| **Prior outpatient visits (2017-2018)** | 12.6 (23.1) | 5.3 (8.5) | 37.2 (36.2) |
| **Prior ED visits (2017-2018)** | 0.8 (3.2) | 0.5 (1.8) | 1.9 (5.6) |
| **Diabetes** | 29,944 (15%) | 18,311 (12%) | 11,633 (25%) |
| **Hypertension** | 62,811 (31%) | 39,169 (25%) | 23,642 (51%) |
| **Heart disease** | 19,780 (9.8%) | 11,216 (7.2%) | 8,564 (19%) |
| **COPD** | 9,652 (4.8%) | 5,707 (3.7%) | 3,945 (8.6%) |
| **Depression** | 36,673 (18%) | 21,973 (14%) | 14,700 (32%) |
| **Obesity** | 36,849 (18%) | 22,327 (14%) | 14,522 (32%) |
| **Cancer** | 19,914 (9.9%) | 10,318 (6.6%) | 9,596 (21%) |
| **BMI** | 29.6 (7.0) | 29.5 (7.0) | 30.1 (6.9) |
| **Household income level** |  |  |  |
| High | 38,247 (19%) | 28,545 (18%) | 9,702 (21%) |
| Low | 74,311 (37%) | 58,470 (38%) | 15,841 (34%) |
| Middle | 50,522 (25%) | 38,852 (25%) | 11,670 (25%) |
| Unknown | 38,136 (19%) | 29,366 (19%) | 8,770 (19%) |
| **Housing instability** | 36,904 (18%) | 29,254 (19%) | 7,650 (17%) |
| **Insurance type** |  |  |  |
| Medicaid | 39,854 (20%) | 31,099 (20%) | 8,755 (19%) |
| Medicare | 36,767 (18%) | 25,280 (16%) | 11,487 (25%) |
| None | 1,326 (0.7%) | 1,179 (0.8%) | 147 (0.3%) |
| Private | 64,800 (32%) | 50,010 (32%) | 14,790 (32%) |
| Unknown | 53,450 (27%) | 44,776 (29%) | 8,674 (19%) |
| VA | 5,019 (2.5%) | 2,889 (1.9%) | 2,130 (4.6%) |
| **Ever smoked** | 84,053 (42%) | 63,851 (41%) | 20,202 (44%) |
| **Ever consumed alcohol** | 178,746 (89%) | 137,349 (88%) | 41,397 (90%) |
| **Functional limitations** | 50,650 (25%) | 35,631 (23%) | 15,019 (33%) |
| **Self-reported general health** |  |  |  |
| Excellent | 22,694 (11%) | 18,982 (12%) | 3,712 (8.1%) |
| Fair | 40,258 (20%) | 28,809 (19%) | 11,449 (25%) |
| Good | 68,204 (34%) | 52,440 (34%) | 15,764 (34%) |
| Poor | 10,199 (5.1%) | 7,069 (4.6%) | 3,130 (6.8%) |
| Very Good | 59,861 (30%) | 47,933 (31%) | 11,928 (26%) |

<sup>*1*</sup>Mean (SD); n (%)

Descriptive analysis revealed meaningful differences in the alignment between healthcare utilization and health need across demographic and socioeconomic subgroups (Figure 2). Among White participants, the proportion classified as high utilizers (25%) exceeded the proportion reporting fair or poor general health status (19%), suggesting that utilization broadly reflects or exceeds health need in this group (Figure 2a). In contrast, Black and Hispanic participants showed the opposite pattern, with a higher proportion reporting fair or poor general health status (33% and 32%, respectively) than were classified as high utilizers (22% and 19%, respectively). Asian participants showed approximate alignment between utilization and health need (16% high utilizers, 15% fair or poor general health status). This divergence indicates that prior healthcare utilization understates health need for Black and Hispanic participants relative to White participants, consistent with the access suppression mechanism proposed in prior work [2, 10]. Similar patterns were observed by insurance type (Figure 2b). Medicaid and uninsured participants showed the largest gaps between health need and utilization, with fair or poor health status substantially exceeding high utilizer classification among Medicaid participants (38% vs 22%) and uninsured participants (24% vs 11%). Privately insured participants showed the opposite pattern, with utilization exceeding health need (23% vs 15%). Medicare participants showed approximate alignment between utilization and health need (31% vs 30%). VA participants showed the highest utilization rate of any insurance group (42%), substantially exceeding their fair or poor health status rate (22%), a pattern that may reflect differences in healthcare access or utilization patterns.

**Figure 2:**
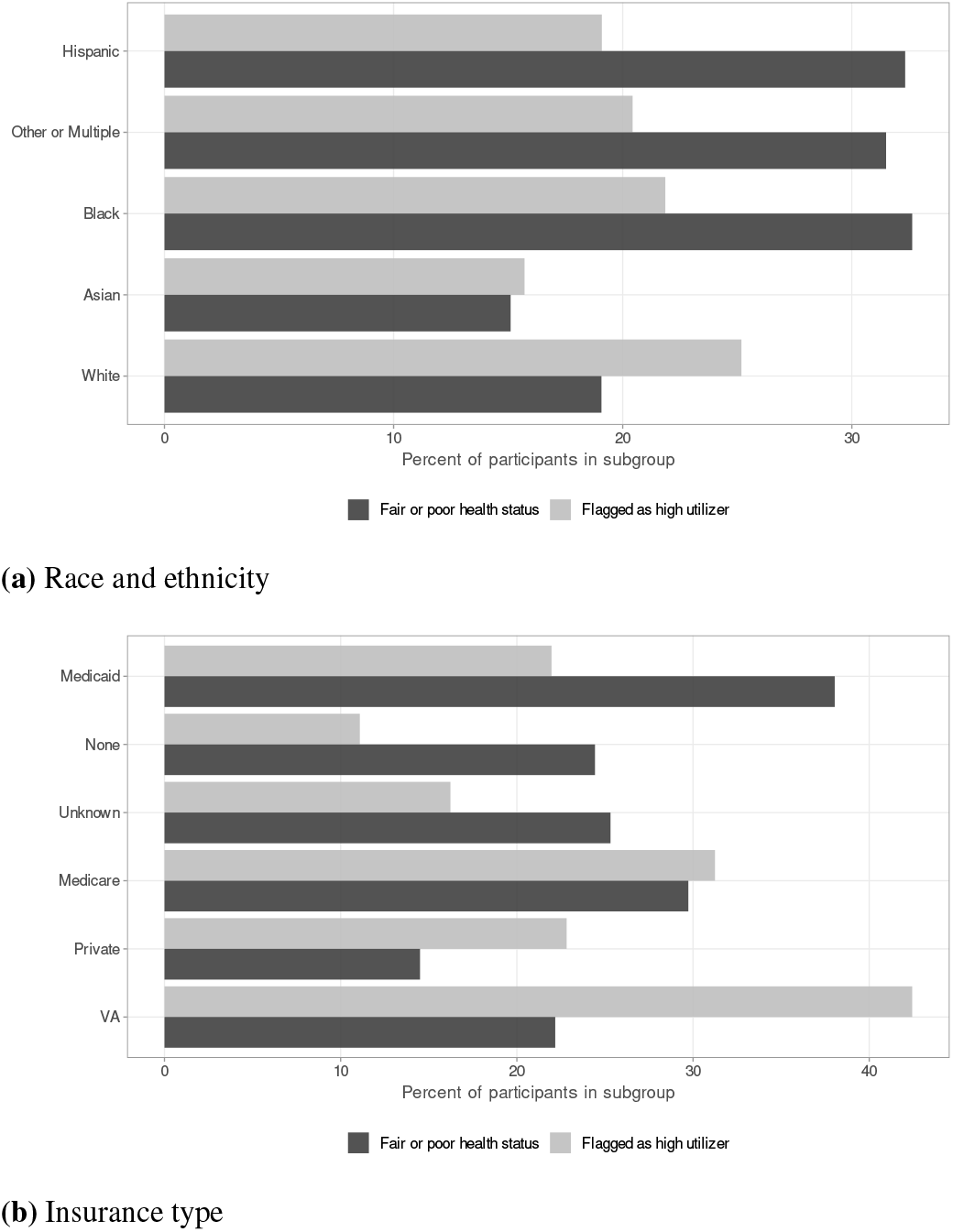
Proportion classified as high utilizers and proportion reporting fair or poor general health status, by (A) race and ethnicity and (B) insurance type. Subgroups in which the proportion reporting fair or poor health exceeds the proportion classified as high utilizers have utilization lower than their reported health burden.

### 3.2. Baseline model performance

All three models demonstrated strong discriminative performance on the held-out test set (Table 2). XGBoost achieved the highest AUC (0.919), followed by random forest (0.916), and logistic regression (0.913), with Brier scores ranging from 0.091 to 0.095 across models. At the 20% selection threshold, overall FNRs were consistent across models, ranging from 0.338 to 0.342, indicating that approximately one in three true high utilizers were not flagged for care management resources, regardless of model class.

**Table 2.**
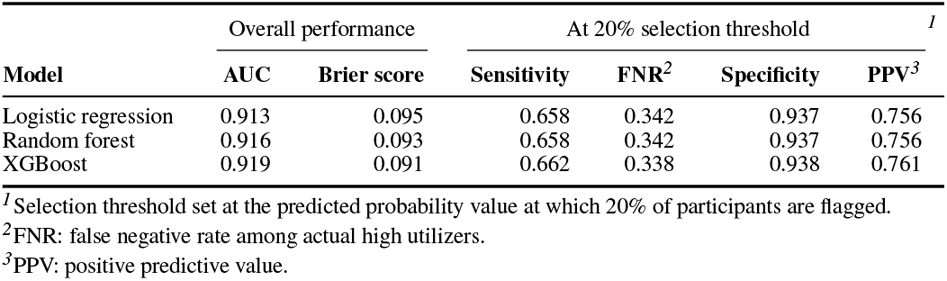
Discrimination and calibration on the held-out test set.

|  | Overall performance |  | At 20% selection threshold<sup>*1*</sup> |  |  |  |
|---|---|---|---|---|---|---|
| **Model** | **AUC** | **Brier score** | **Sensitivity** | **FNR**<sup>*2*</sup> | **Specificity** | **PPV**<sup>*3*</sup> |
| Logistic regression | 0.913 | 0.095 | 0.658 | 0.342 | 0.937 | 0.756 |
| Random forest | 0.916 | 0.093 | 0.658 | 0.342 | 0.937 | 0.756 |
| XGBoost | 0.919 | 0.091 | 0.662 | 0.338 | 0.938 | 0.761 |

<sup>*1*</sup>Selection threshold set at the predicted probability value at which 20% of participants are flagged.
<sup>*2*</sup>FNR: false negative rate among actual high utilizers.
<sup>*3*</sup>PPV: positive predictive value.

### 3.3. Baseline allocation equity

At the 20% threshold, all three models produced consistent patterns of FNR disparity across racial and ethnic groups. Under logistic regression, Black and Hispanic participants showed lower true positive rates than White participants, with EODs of -0.048 and -0.038, respectively, indicating that the model correctly identified a smaller proportion of high utilizers from these groups relative to White participants. Asian participants showed the largest disparity, with an EOD of -0.124 under logistic regression and -0.112 under XGBoost.

The allocation rate conditional on need, or the proportion of fair or poor health participants flagged for care management resources, revealed a similar pattern. Among participants with fair or poor health status, White participants were flagged at the highest rate (29.5% under logistic regression), while Asian participants were flagged at the lowest rate (21.4%), representing a 8.1 percentage point gap. Black and Hispanic participants were flagged at rates of 25.7% and 25.2%, respectively, gaps of 3.8 and 4.3 percentage points relative to White participants. The Asian allocation rate gap was somewhat smaller under random forest (6.6 percentage points) compared to logistic regression (8.1 percentage points) and XGBoost (7.8 percentage points), though the direction of disparity was consistent across all model classes.

Disparities by insurance type were more pronounced. Uninsured participants showed the largest FNR disparity of any subgroup, with an EOD of -0.148 under logistic regression, indicating that the model’s true positive rate among uninsured high utilizers was 14.8 percentage points below that of privately insured high utilizers. Medicaid participants showed a smaller but meaningful disparity (EOD -0.034). In contrast, Medicare and VA participants showed positive EOD values (0.100 and 0.182, respectively), indicating lower FNR than privately insured participants.

The allocation rate conditional on need by insurance type reinforced these findings. Uninsured participants with fair or poor health status were flagged at a rate of 13.9%, compared to 27.9% for privately insured participants, a gap of 14.0 percentage points representing the largest allocation disparity for any subgroup. Medicaid participants were flagged at 23.4%, a gap of 4.5 percentage points relative to Private. VA participants showed the highest allocation rate of any insurance group (48.2%).

### 3.4. Disparity decomposition

The endowments component accounted for nearly all of the Black-White gap in predicted allocation log-odds (0.211 of 0.212 total), with negligible coefficients (-0.0003) and interaction (0.001) components.

Prior outpatient visits contributed the largest single endowment (0.215 log-odds units), slightly exceeding the total endowments component and indicating that outpatient utilization is the dominant characteristic difference contributing to the Black-White gap in predicted allocation (Figure 3). All remaining predictors contributed a combined endowment of -0.004, partially offsetting the outpatient difference. The largest negative contributions came from prior emergency department visits, reflecting higher average emergency department utilization among Black participants.

**Figure 3:**
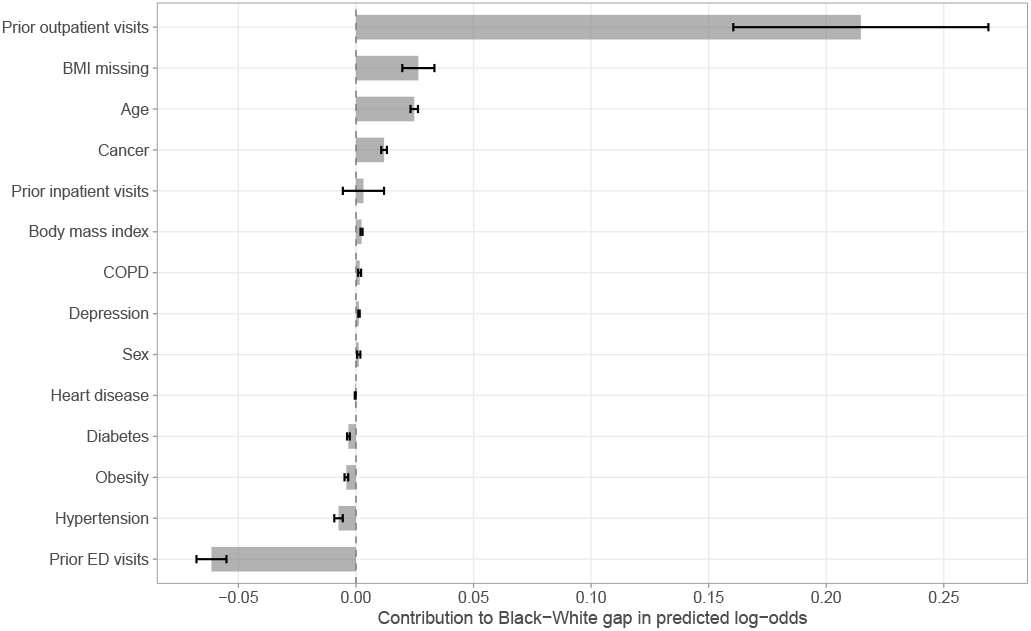
Contribution of each predictor to the Black–White gap in predicted log-odds. Error bars show 95% bootstrap confidence intervals.

### 3.5. Simulation 1: threshold variation

Varying the selection threshold produced modest and variable effects on equity outcomes across subgroups and model classes.

#### 3.5.1. False Negative Rate and Equal Opportunity Difference

FNR disparities persisted across all threshold conditions (Table 3). Under logistic regression, Asian participants showed the largest and most persistent FNR disparity across all threshold conditions (EOD: -0.103 at 10% threshold, -0.124 at 20% threshold, -0.098 at 30% threshold). Black participants showed increasing FNR disparity from the 10% to 20% threshold (EOD: -0.019 to -0.048) with no further change at 30%, while Hispanic participants showed gradual worsening across all three thresholds (EOD: -0.027, -0.039, -0.051). The direction of change in racial EOD across threshold conditions differed by model class, with logistic regression showing a modest worsening while random forest and XGBoost showed a slight narrowing as the selection threshold increased (Figure 4a).

**Table 3.** False negative rate and equal opportunity difference across selection threshold conditions, by race/ethnicity and insurance type.

|  | 10% threshold |  | 20% threshold |  | 30% threshold |  |
|---|---|---|---|---|---|---|
| Subgroup<sup>*1*</sup> | FNR | EOD<sup>*2*</sup> | FNR | EOD<sup>*2*</sup> | FNR | EOD<sup>*2*</sup> |
| **Race/ethnicity** |  |  |  |  |  |  |
| White | 0.600 | 0.000 | 0.322 | 0.000 | 0.170 | 0.000 |
| Hispanic | 0.627 | -0.027 | 0.362 | -0.039 | 0.221 | -0.051 |
| Other or Multiple | 0.626 | -0.026 | 0.362 | -0.040 | 0.218 | -0.047 |
| Black | 0.620 | -0.019 | 0.371 | -0.048 | 0.219 | -0.048 |
| Asian | 0.704 | -0.103 | 0.446 | -0.124 | 0.268 | -0.098 |
| **Insurance type** |  |  |  |  |  |  |
| VA | 0.451 | 0.204 | 0.192 | 0.182 | 0.092 | 0.115 |
| Medicare | 0.560 | 0.094 | 0.274 | 0.100 | 0.140 | 0.068 |
| Private | 0.654 | 0.000 | 0.374 | 0.000 | 0.207 | 0.000 |
| Medicaid | 0.649 | 0.006 | 0.408 | -0.034 | 0.258 | -0.051 |
| None | 0.674 | -0.019 | 0.522 | -0.148 | 0.370 | -0.162 |

<sup>*1*</sup>Reference groups: White for race/ethnicity, private insurance for insurance type.
<sup>*2*</sup>EOD computed relative to reference group.

**Figure 4:**
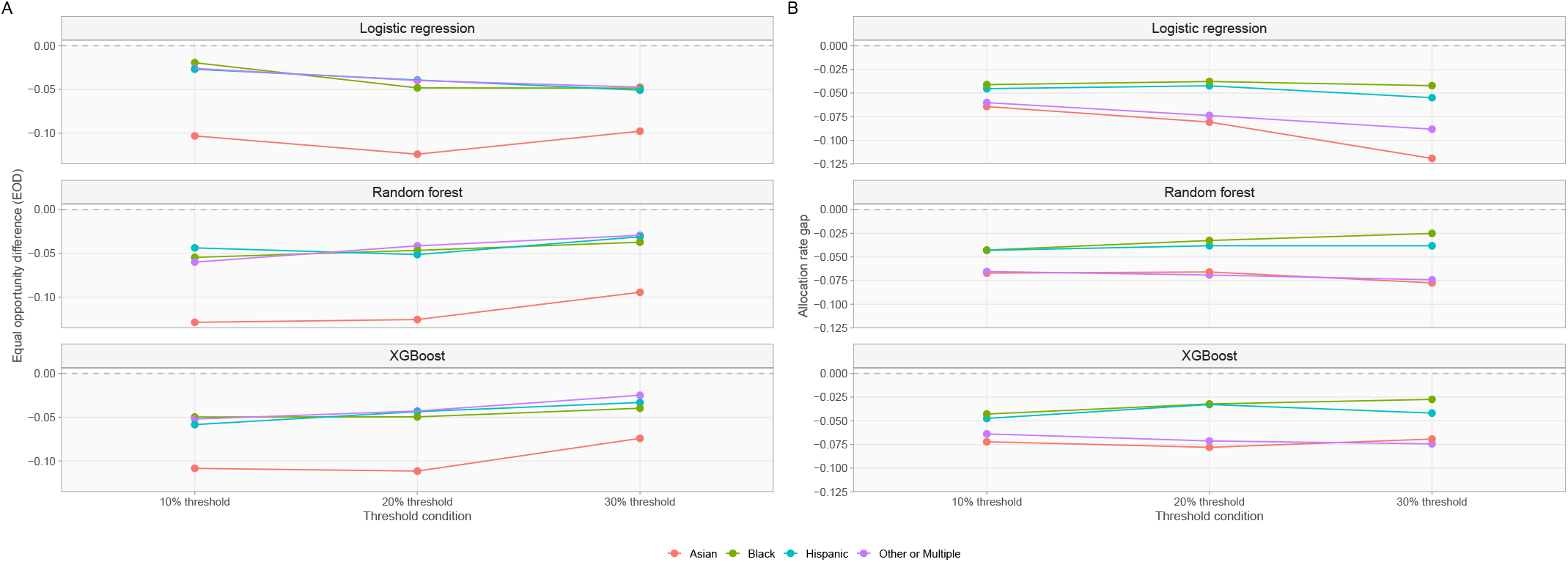
Equal opportunity difference (A) and allocation rate gap (B) across selection thresholds, by race and ethnicity. Estimates are relative to White participants.

By insurance type, uninsured participants showed the strongest and most consistent threshold-dependent worsening across all three model classes (logistic regression EOD: -0.019 at 10% threshold, -0.148 at 20% threshold, -0.162 at 30% threshold). Medicaid participants showed near-zero FNR disparity at the 10% threshold (EOD: 0.006) then worsened at higher threshold conditions (-0.034 at 20% threshold, -0.051 at 30% threshold). VA and Medicare participants showed consistently positive EOD across all threshold conditions and model classes, indicating lower FNR than privately insured participants regardless of threshold (Figure 5a).

**Figure 5:**
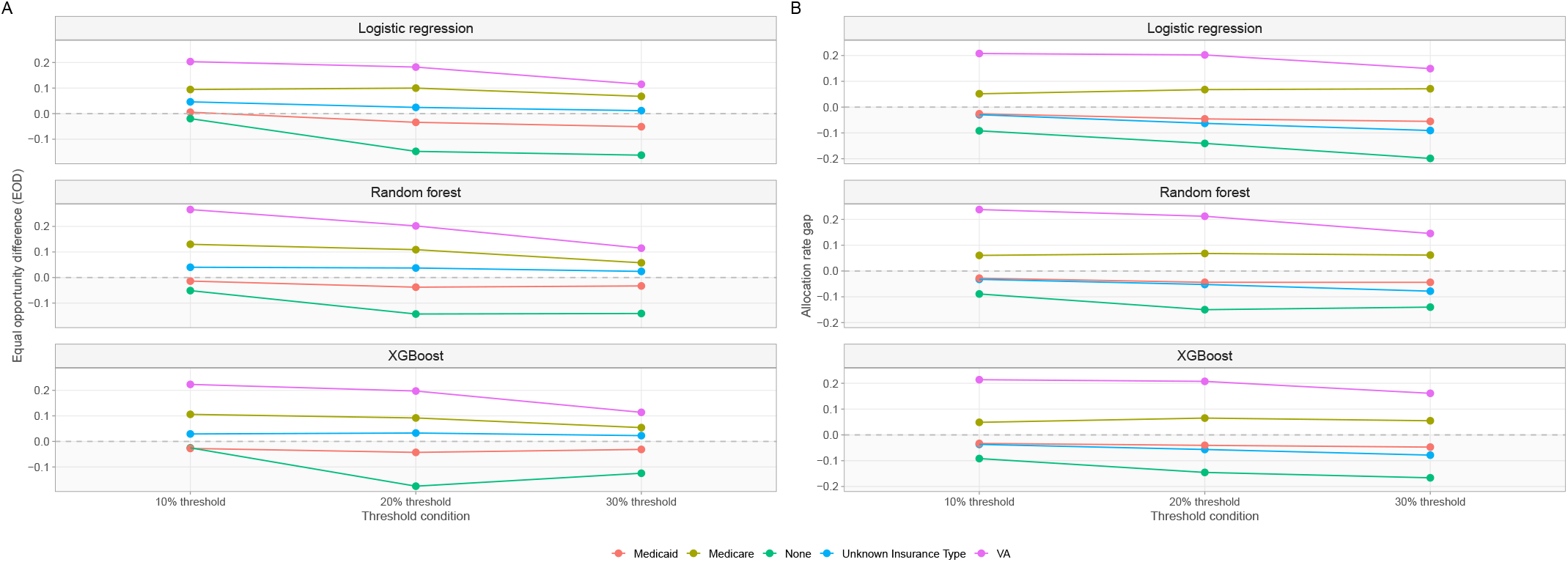
Equal opportunity difference (A) and allocation rate gap (B) across selection thresholds, by insurance type. Estimates are relative to private insurance.

#### 3.5.2. Allocation Rate Conditional on Need

Allocation rate conditional on need showed persistent gaps across all threshold conditions and racial and ethnic groups (Table 4). Under logistic regression, White participants were flagged at the highest rate among fair or poor health participants at every threshold condition (10% threshold: 17.9%, 20% threshold: 29.5%, 30% threshold: 40.7%), while Asian participants were consistently flagged at the lowest rate. The Black-White and Hispanic-White allocation gaps remained relatively stable across threshold conditions (Black: -4.1, -3.8, -4.2 percentage points; Hispanic: -4.5, -4.2, -5.5 percentage points), while the Asian-White gap widened at higher thresholds (-6.4, -8.1, -11.9 percentage points) (Figure 4b).

**Table 4.**
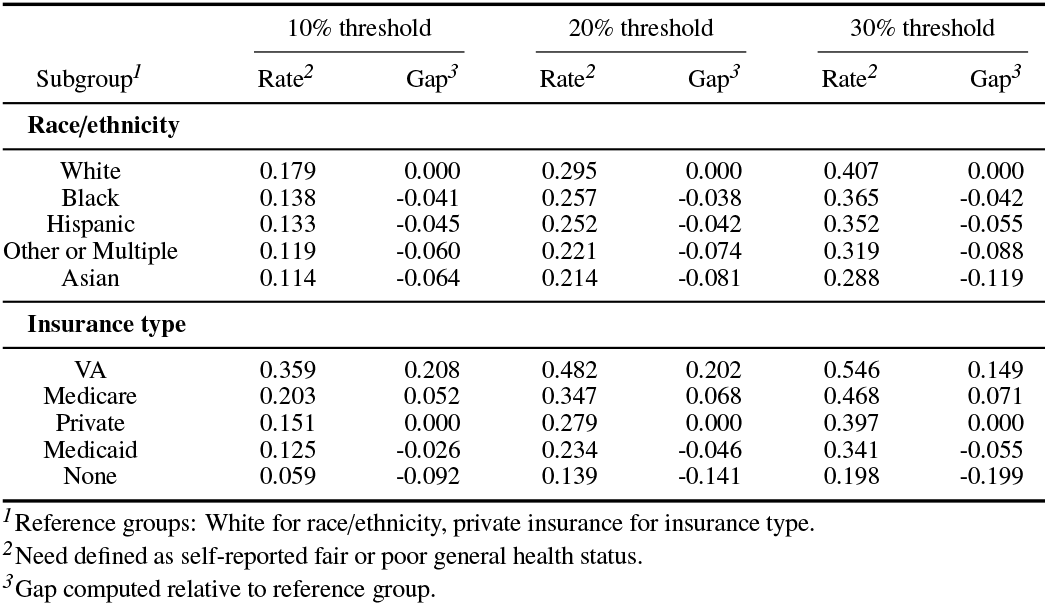
Allocation rate conditional on need and allocation rate gap across selection threshold conditions, by race/ethnicity and insurance type.

Allocation rate gaps by insurance type were relatively stable across threshold conditions for most groups. Uninsured participants showed the most notable widening gap across thresholds (-9.2, -14.1, -19.9 percentage points) relative to privately insured participants, with uninsured participants flagged at less than half the rate of privately insured participants at the 30% threshold (19.8% vs 39.7%). VA participants maintained the highest allocation rate of any insurance group across all threshold conditions (Figure 5b).

### 3.6. Simulation 2: feature set variation

Feature set variation produced mixed and group-specific effects on both model accuracy and equity outcomes.

#### 3.6.1. Model Discrimination

Removing all prior utilization variables reduced model discrimination across all three model classes (AUC: 0.739-0.749, compared to 0.913-0.919 under standard). Removing only prior outpatient visits partially recovered predictive performance (AUC: 0.774-0.784), while the survey-enriched feature set maintained accuracy equivalent to the standard feature set across all model classes (AUC: 0.913-0.920) (Table 5).

**Table 5.** Discrimination across predictor feature set conditions.

|  | Standard<sup>*1*</sup> | Full utilization restricted<sup>*2*</sup> | Outpatient restricted<sup>*3*</sup> | Survey-enriched<sup>*4*</sup> |
|---|---|---|---|---|
| **Logistic regression** | 0.913 | 0.739 | 0.774 | 0.913 |
| **Random forest** | 0.916 | 0.740 | 0.780 | 0.918 |
| **XGBoost** | 0.919 | 0.742 | 0.784 | 0.920 |

<sup>*1*</sup>Age, sex, prior inpatient visits, prior emergency department visits, prior outpatient visits, diabetes, hypertension, heart disease, chronic obstructive pulmonary disease, depression, obesity, cancer, chronic condition count, body mass index, and body mass index missing indicator.
<sup>*2*</sup>Standard feature set excluding prior inpatient, emergency department, and outpatient visits.
<sup>*3*</sup>Standard feature set excluding prior outpatient visits.
<sup>*4*</sup>Standard feature set plus household income level and missing indicator, insurance type, housing instability, ever smoked, ever used alcohol, and functional limitations.

#### 3.6.2. False Negative Rate and Equal Opportunity Difference

Under the full utilization restricted feature set, FNR disparities worsened for Asian participants (EOD: -0.209, compared to -0.124 under the standard feature set), while Black participants showed a modest narrowing (EOD: -0.034 vs -0.048) (Table 6). Under the outpatient restricted feature set, Black participants achieved near-zero EOD under logistic regression (0.001), approaching FNR parity with White participants. This pattern is consistent with the Oaxaca-Blinder decomposition finding that prior outpatient utilization was the primary characteristic difference contributing to the Black-White allocation gap. However, FNR for high utilization rose in both groups, so the improvements reflect convergence rather than better identification of Black high utilizers (White FNR: 0.322 to 0.538; Black FNR: 0.371 to 0.536). The survey-enriched feature set produced a modest but consistent worsening of FNR disparities for Black, Hispanic, and Other or Multiple participants relative to standard (Black EOD: -0.065 vs -0.048; Hispanic EOD: -0.055 vs -0.039; Other or Multiple EOD: -0.051 vs -0.040), while Asian participants showed slight improvement (EOD: - 0.110 vs -0.124) (Figure 6a).

**Table 6.** False negative rate and equal opportunity difference across predictor feature set conditions, by race/ethnicity and insurance type.

| Subgroup<sup>*5*</sup> | Standard<sup>*1*</sup> |  | Full utilization restricted<sup>*2*</sup> |  | Outpatient restricted<sup>*3*</sup> |  | Survey-enriched<sup>*4*</sup> |  |
|---|---|---|---|---|---|---|---|---|
|  | FNR | EOD<sup>*6*</sup> | FNR | EOD<sup>*6*</sup> | FNR | EOD<sup>*6*</sup> | FNR | EOD<sup>*6*</sup> |
| **Race/ethnicity** |  |  |  |  |  |  |  |  |
| White | 0.322 | 0.000 | 0.576 | 0.000 | 0.538 | 0.000 | 0.315 | 0.000 |
| Hispanic | 0.362 | -0.039 | 0.623 | -0.047 | 0.594 | -0.056 | 0.370 | -0.055 |
| Other or Multiple | 0.362 | -0.040 | 0.617 | -0.041 | 0.585 | -0.048 | 0.366 | -0.051 |
| Black | 0.371 | -0.048 | 0.610 | -0.034 | 0.536 | 0.001 | 0.380 | -0.065 |
| Asian | 0.446 | -0.124 | 0.786 | -0.209 | 0.739 | -0.201 | 0.425 | -0.110 |
| **Insurance type** |  |  |  |  |  |  |  |  |
| VA | 0.192 | 0.182 | 0.539 | 0.132 | 0.540 | 0.083 | 0.147 | 0.208 |
| Medicare | 0.274 | 0.100 | 0.445 | 0.226 | 0.413 | 0.210 | 0.265 | 0.091 |
| Private | 0.374 | 0.000 | 0.671 | 0.000 | 0.623 | 0.000 | 0.356 | 0.000 |
| Medicaid | 0.408 | -0.034 | 0.654 | 0.017 | 0.584 | 0.039 | 0.401 | -0.046 |
| None | 0.522 | -0.148 | 0.804 | -0.133 | 0.783 | -0.160 | 0.587 | -0.231 |

<sup>*1*</sup>Age, sex, prior inpatient visits, prior emergency department visits, prior outpatient visits, diabetes, hypertension, heart disease, chronic obstructive pulmonary disease, depression, obesity, cancer, chronic condition count, body mass index, and body mass index missing indicator.
<sup>*2*</sup>Standard feature set excluding prior inpatient, emergency department, and outpatient visits.
<sup>*3*</sup>Standard feature set excluding prior outpatient visits.
<sup>*4*</sup>Standard feature set plus household income level and missing indicator, insurance type, housing instability, ever smoked, ever used alcohol, and functional limitations.
<sup>*5*</sup>Reference groups: White for race/ethnicity, private insurance for insurance type.
<sup>*6*</sup>EOD computed relative to reference group.

**Figure 6:**
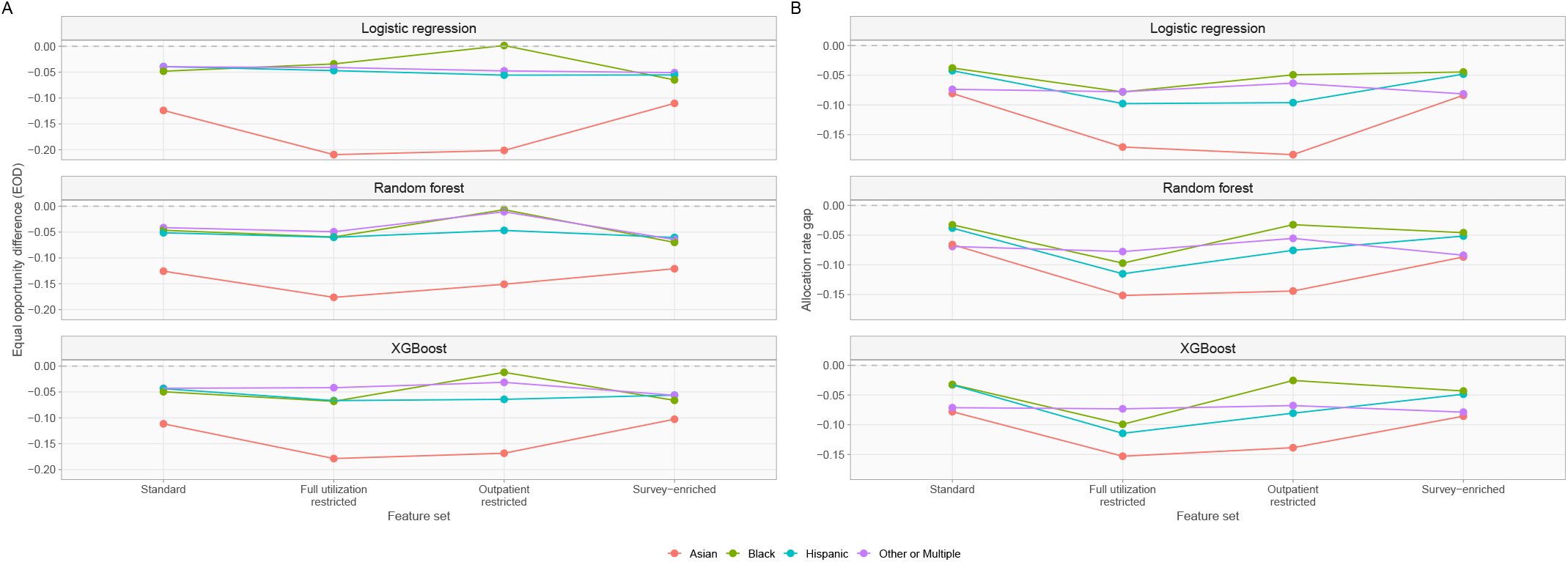
Equal opportunity difference (A) and allocation rate gap (B) across feature sets, by race and ethnicity. Estimates are relative to White participants.

By insurance type, uninsured participants showed the largest FNR disparity across all feature set conditions, worsening under the survey-enriched feature set (EOD: -0.231 vs -0.148 under standard). Under the outpatient restricted feature set, uninsured EOD showed opposite patterns by model class, worsening under logistic regression (-0.160) but improving under random forest (-0.078) and XGBoost (-0.131). Medicaid participants showed a notable reversal across feature set conditions, with EOD shifting from negative under standard (-0.034) to positive under both restricted conditions (full utilization restricted: 0.017, outpatient restricted: 0.039). VA and Medicare participants maintained positive EOD across all feature set conditions and model classes (Figure 7a).

**Figure 7:**
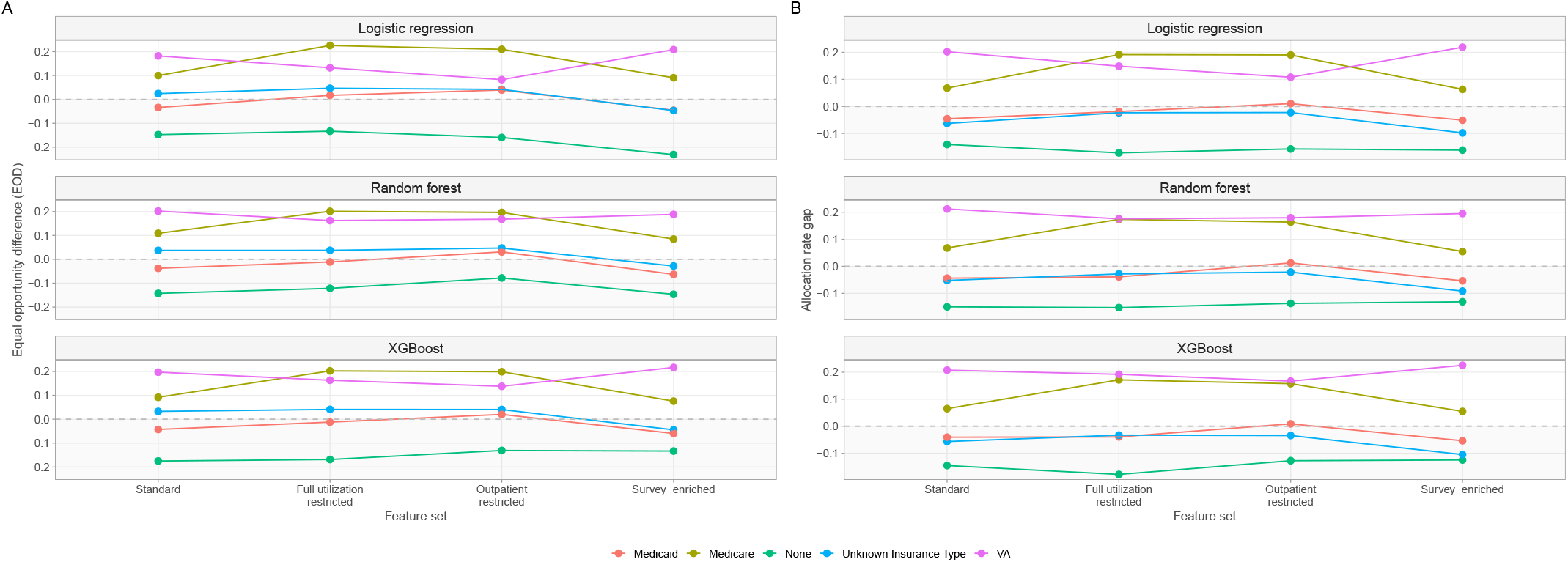
Equal opportunity difference (A) and allocation rate gap (B) across feature sets, by insurance type. Estimates are relative to private insurance.

**Figure 8:**
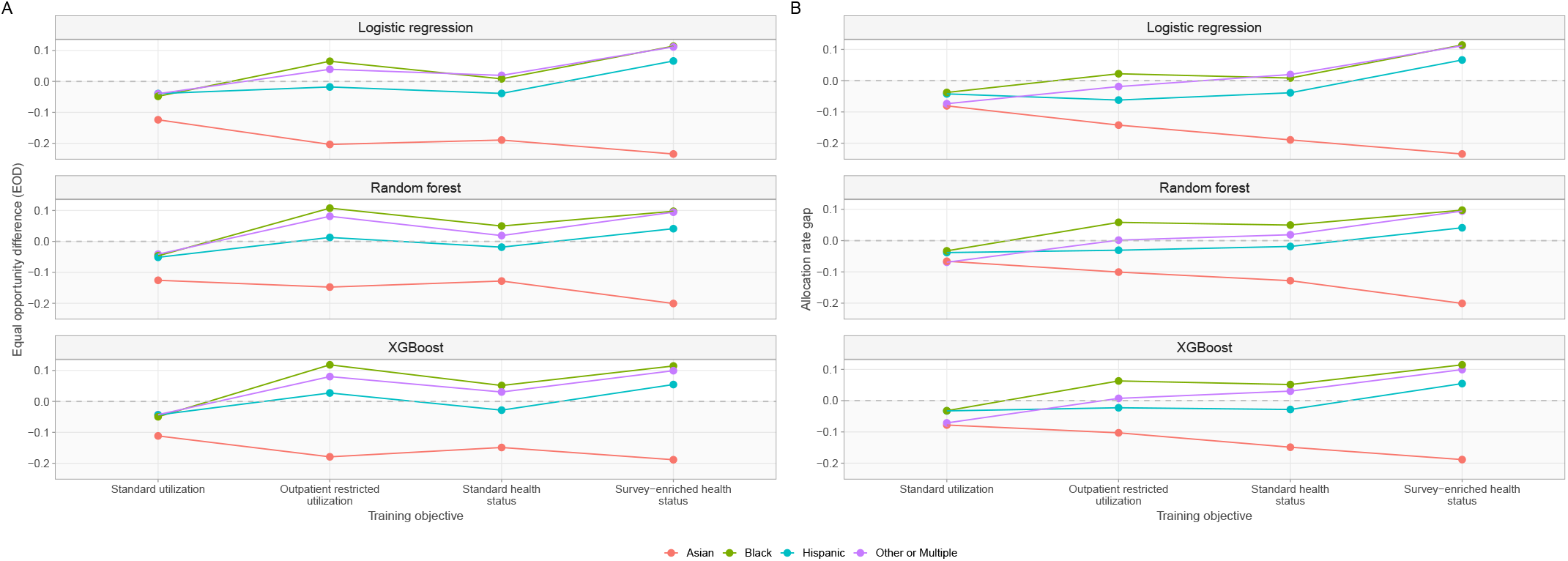
Equal opportunity difference (A) and allocation rate gap (B) across training objectives, by race and ethnicity. Estimates are relative to White participants.

**Figure 9:**
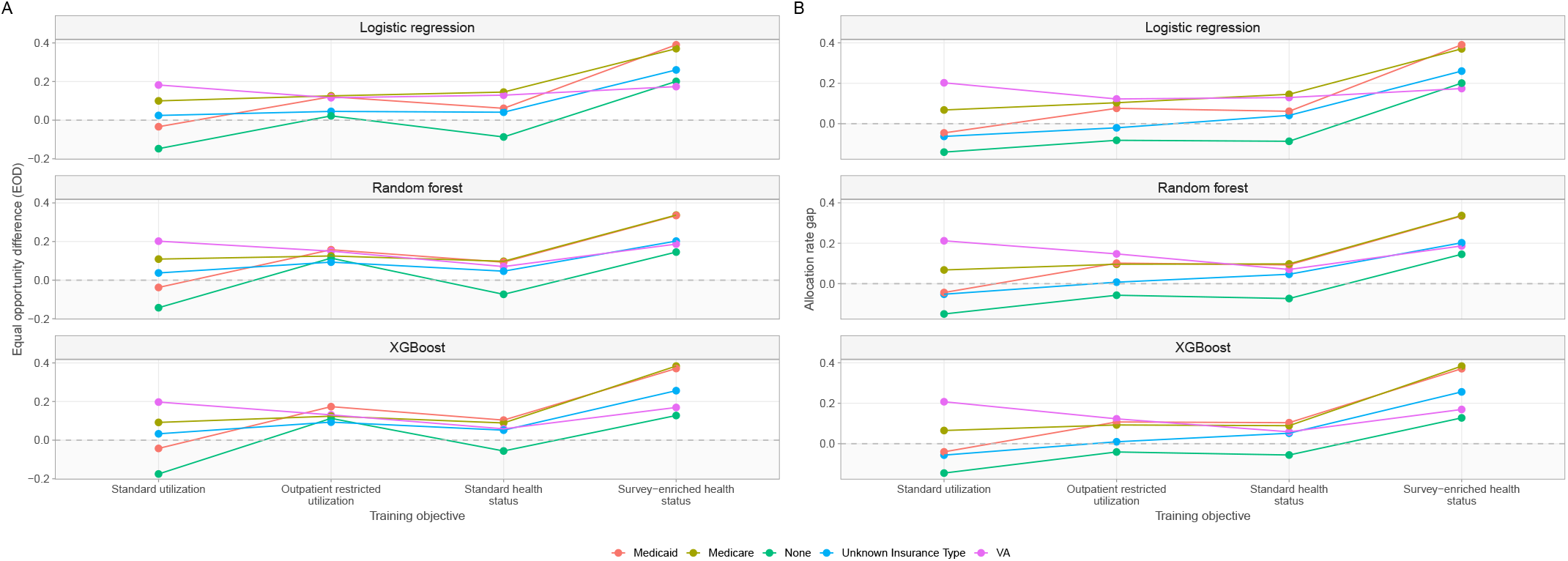
Equal opportunity difference (A) and allocation rate gap (B) across training objectives, by insurance type. Estimates are relative to private insurance.

**Figure 10:**
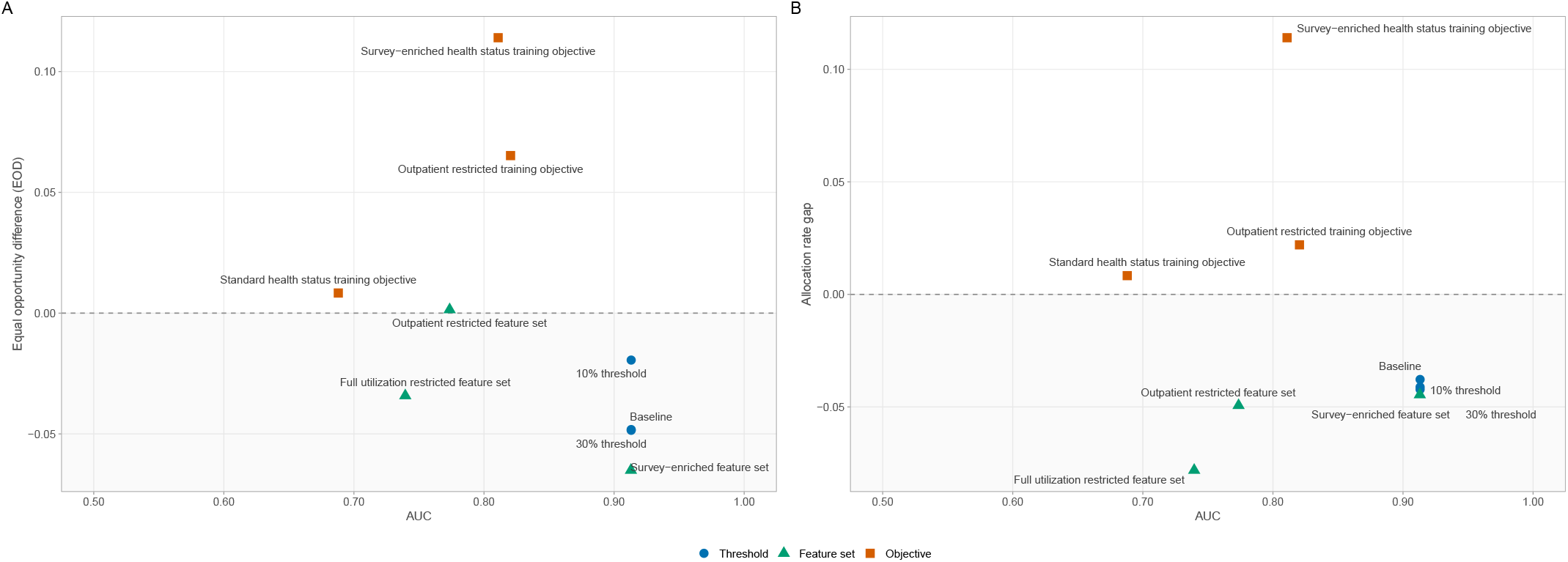
Equal opportunity difference (A) and allocation rate gap (B) against AUC for Black participants relative to White participants, across all simulation conditions. Shaded regions indicate conditions in which Black participants were disadvantaged relative to the reference group. Estimates shown for logistic regression.

**Figure 11:**
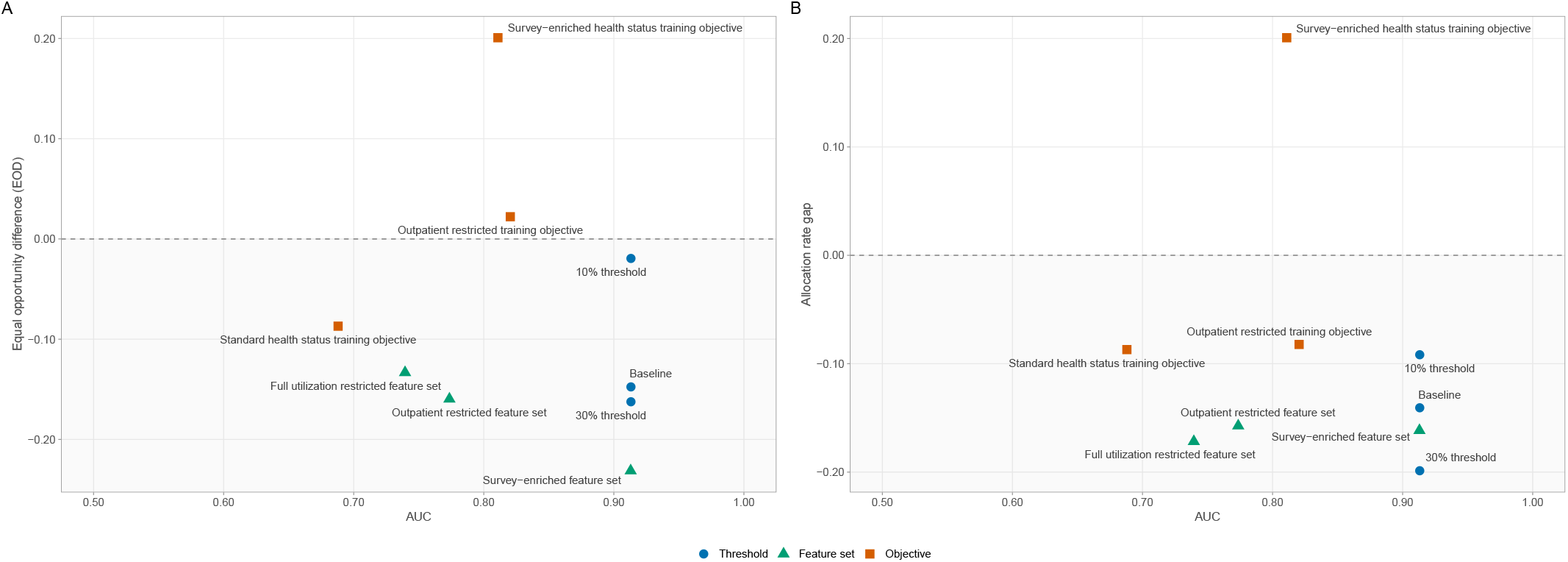
Equal opportunity difference (A) and allocation rate gap (B) against AUC for uninsured participants relative to privately insured participants, across all simulation conditions. Shaded regions indicate conditions in which uninsured participants were disadvantaged relative to the reference group. Estimates shown for logistic regression.

#### 3.6.3. Allocation Rate Conditional on Need

Racial allocation rate gaps were generally stable under the survey-enriched feature set relative to standard, while both restricted conditions produced notable changes. The Asian-White allocation rate gap worsened under both restricted conditions (full utilization restricted: -17.1 percentage points, outpatient restricted: -18.4 percentage points, compared to -8.1 percentage points under standard), representing the largest racial allocation rate disparity across all Simulation 2 conditions (Table 7). Hispanic-White gaps also worsened under both restricted conditions (full utilization restricted: -9.8 percentage points, outpatient restricted: -9.6 percentage points, compared to -4.2 percentage points under standard). Black participants showed a different pattern, with the full utilization restricted condition worsening the Black-White gap (-7.8 percentage points vs -3.8 under standard), while the outpatient restricted condition partially recovered it (-4.9 percentage points) (Figure 4b).

**Table 7.** Allocation rate conditional on need and allocation rate gap across predictor feature set conditions, by race/ethnicity and insurance type.

|  | Standard<sup>*1*</sup> |  | Full utilization restricted<sup>*2*</sup> |  | Outpatient restricted<sup>*3*</sup> |  | Survey-enriched<sup>*4*</sup> |  |
|---|---|---|---|---|---|---|---|---|
| Subgroup<sup>*5*</sup> | Rate<sup>*6*</sup> | Gap<sup>*7*</sup> | Rate<sup>*6*</sup> | Gap<sup>*7*</sup> | Rate<sup>*6*</sup> | Gap<sup>*7*</sup> | Rate<sup>*6*</sup> | Gap<sup>*7*</sup> |
| **Race/ethnicity** |  |  |  |  |  |  |  |  |
| White | 0.295 | 0.000 | 0.366 | 0.000 | 0.350 | 0.000 | 0.298 | 0.000 |
| Black | 0.257 | -0.038 | 0.288 | -0.078 | 0.300 | -0.049 | 0.253 | -0.045 |
| Hispanic | 0.252 | -0.042 | 0.269 | -0.098 | 0.253 | -0.096 | 0.250 | -0.048 |
| Other or Multiple | 0.221 | -0.074 | 0.289 | -0.078 | 0.286 | -0.063 | 0.216 | -0.081 |
| Asian | 0.214 | -0.081 | 0.196 | -0.171 | 0.166 | -0.184 | 0.214 | -0.084 |
| **Insurance type** |  |  |  |  |  |  |  |  |
| VA | 0.482 | 0.202 | 0.429 | 0.149 | 0.374 | 0.108 | 0.509 | 0.219 |
| Medicare | 0.347 | 0.068 | 0.472 | 0.192 | 0.457 | 0.190 | 0.353 | 0.063 |
| Private | 0.279 | 0.000 | 0.281 | 0.000 | 0.266 | 0.000 | 0.290 | 0.000 |
| Medicaid | 0.234 | -0.046 | 0.262 | -0.019 | 0.277 | 0.010 | 0.239 | -0.051 |
| None | 0.139 | -0.141 | 0.109 | -0.172 | 0.109 | -0.157 | 0.129 | -0.162 |

<sup>*1*</sup>Age, sex, prior inpatient visits, prior emergency department visits, prior outpatient visits, diabetes, hypertension, heart disease, chronic obstructive pulmonary disease, depression, obesity, cancer, chronic condition count, body mass index, and body mass index missing indicator.
<sup>*2*</sup>Standard feature set excluding prior inpatient, emergency department, and outpatient visits.
<sup>*3*</sup>Standard feature set excluding prior outpatient visits.
<sup>*4*</sup>Standard feature set plus household income level and missing indicator, insurance type, housing instability, ever smoked, ever used alcohol, and functional limitations.
<sup>*5*</sup>Reference groups: White for race/ethnicity, private insurance for insurance type.
<sup>*6*</sup>Need defined as self-reported fair or poor general health status.
<sup>*7*</sup>Gap computed relative to reference group.

By insurance type, uninsured participants showed persistently large allocation rate gaps across all feature set conditions (-14.1 to -17.2 percentage points), with no feature set meaningfully closing this disparity. Medicaid participants approached allocation rate parity under the outpatient restricted feature set (1.0 percentage point gap compared to privately insured participants). Medicare participants showed higher allocation rates than privately insured participants under both restricted conditions (19.0-19.2 percentage point gap) (Figure 5b).

### 3.7. Simulation 3: objective variation

Training objective variation produced substantial changes in equity outcomes across subgroups, with the direction of disparities shifting across conditions.

#### 3.7.1. Model Discrimination

The standard utilization objective maintained the highest AUC across all model classes (Table 8). The outpatient restricted utilization objective moderately reduced discrimination ability (logistic regression: 0.820, random forest: 0.837, XGBoost: 0.839), reflecting the narrower outcome definition based on inpatient and emergency department visits only. The standard health status objective produced the largest reduction in discrimination across all model classes (logistic regression: 0.688, random forest: 0.723, XGBoost: 0.725), reflecting the greater difficulty of predicting self-reported general health status from prior utilization and clinical characteristics. Adding socioeconomic predictors under the survey-enriched health status objective recovered discrimination ability (logistic regression: 0.811, random forest: 0.819, XGBoost: 0.822), approaching the accuracy of the outpatient restricted utilization objective.

**Table 8.** Discrimination across training objective conditions.

|  | Standard utilization<sup>*1*</sup> | Outpatient restricted utilization<sup>*2*</sup> | Standard health status<sup>*3*</sup> | Survey-enriched health status<sup>*4*</sup> |
|---|---|---|---|---|
| **Logistic regression** | 0.913 | 0.820 | 0.688 | 0.811 |
| **Random forest** | 0.916 | 0.837 | 0.723 | 0.819 |
| **XGBoost** | 0.919 | 0.839 | 0.725 | 0.822 |

<sup>*1*</sup>High utilization based on inpatient, emergency department, and outpatient visit count, using the standard feature set.
<sup>*2*</sup>High utilization based on inpatient and emergency department visit count, using the standard feature set.
<sup>*3*</sup>Self-reported fair or poor general health status, using the standard feature set.
<sup>*4*</sup>Self-reported fair or poor general health status, using the survey-enriched feature set.

#### 3.7.2. False Negative Rate and Equal Opportunity Difference

Under the outpatient restricted utilization objective, Black participants showed a notable reversal in EOD relative to standard utilization (0.065 vs -0.048), indicating lower FNR than White participants (Table 9). Hispanic EOD narrowed (-0.018 vs -0.039), while Asian participants showed further worsening (-0.204 vs -0.124). Under the standard health status objective, racial EOD gaps narrowed for most groups, with Black EOD approaching zero (0.008) and Other or Multiple participants showing slightly positive (0.020). Hispanic EOD remained similar to standard utilization (-0.039), and Asian participants continued to show the largest negative EOD (-0.189). The surveyenriched health status objective reversed the direction of disparities for most racial groups, with Black, Hispanic, and Other or Multiple participants all showing positive EOD (0.114, 0.066, 0.111, respectively), indicating lower FNR than White participants. Asian participants remained the exception across all objective conditions, showing consistently negative and worsening EOD (outpatient restricted utilization: -0.204, standard health status: -0.189, survey-enriched health status: -0.235).

**Table 9.** False negative rate and equal opportunity difference across training objective conditions, by race/ethnicity and insurance type.

| Subgroup<sup>*5*</sup> | Standard utilization<sup>*1*</sup> |  | Outpatient restricted utilization<sup>*2*</sup> |  | Standard health status<sup>*3*</sup> |  | Survey-enriched health status<sup>*4*</sup> |  |
|---|---|---|---|---|---|---|---|---|
|  | FNR | EOD | FNR | EOD | FNR | EOD | FNR | EOD |
| **Race/ethnicity** |  |  |  |  |  |  |  |  |
| White | 0.322 | 0.000 | 0.457 | 0.000 | 0.604 | 0.000 | 0.559 | 0.000 |
| Hispanic | 0.362 | -0.039 | 0.475 | -0.018 | 0.643 | -0.039 | 0.493 | 0.066 |
| Other or Multiple | 0.362 | -0.040 | 0.418 | 0.039 | 0.584 | 0.020 | 0.447 | 0.111 |
| Black | 0.371 | -0.048 | 0.392 | 0.065 | 0.596 | 0.008 | 0.445 | 0.114 |
| Asian | 0.446 | -0.124 | 0.661 | -0.204 | 0.793 | -0.189 | 0.793 | -0.235 |
| **Insurance type** |  |  |  |  |  |  |  |  |
| VA | 0.192 | 0.182 | 0.398 | 0.116 | 0.546 | 0.129 | 0.601 | 0.174 |
| Medicare | 0.274 | 0.100 | 0.388 | 0.126 | 0.530 | 0.145 | 0.405 | 0.370 |
| Private | 0.374 | 0.000 | 0.514 | 0.000 | 0.675 | 0.000 | 0.775 | 0.000 |
| Medicaid | 0.408 | -0.034 | 0.392 | 0.121 | 0.614 | 0.061 | 0.385 | 0.390 |
| None | 0.522 | -0.148 | 0.492 | 0.022 | 0.762 | -0.087 | 0.574 | 0.201 |

Note: False negative rate is computed against each condition’s own outcome definition and is therefore not directly comparable across objective conditions.
<sup>*1*</sup>High utilization based on inpatient, emergency department, and outpatient visit count, using the standard feature set.
<sup>*2*</sup>High utilization based on inpatient and emergency department visit count, using the standard feature set.
<sup>*3*</sup>Self-reported fair or poor general health status, using the standard feature set.
<sup>*4*</sup>Self-reported fair or poor general health status, using the survey-enriched feature set.
<sup>*5*</sup>Reference groups: White for race/ethnicity, private insurance for insurance type.

By insurance type, uninsured participants showed the largest shifts across objective conditions. Under standard utilization, uninsured participants showed the largest negative EOD of any insurance group (-0.148), which narrowed to near-zero under outpatient restricted utilization (0.022), remained negative under standard health status (-0.087), and reversed under surveyenriched health status (0.201). Medicaid participants showed a similar pattern under the survey-enriched health status condition (standard utilization: -0.034, outpatient restricted utilization: 0.121, standard health status: 0.061, survey-enriched health status: 0.390). VA and Medicare maintained positive EOD across all objective conditions, with Medicare showing a particularly large positive EOD under survey-enriched health status (0.370).

#### 3.7.3. Allocation Rate Conditional on Need

For health status objective conditions, the allocation rate conditional on need and EOD are equivalent metrics, as the model outcome and equity benchmark are both defined by self-reported general health status.

Under the outpatient restricted utilization objective, Black participant allocation rates increased relative to standard utilization (34.5% vs 25.7%), with the Black-White gap reversing (+2.2 percentage points vs -3.8 under standard utilization) (Table 10). Hispanic allocation rates showed little change (26.1% vs 25.2%), with the gap slightly worsening (-6.2 vs - 4.2 percentage points). Asian allocation rates declined (18.1% vs 21.4%), with the gap worsening (-14.2 percentage points). Under the standard health status objective, most racial groups showed allocation rates closer to the White reference (39.6%) than under standard utilization. Black (40.4%), Other or Multiple (41.6%), and Hispanic (35.7%) participants showed positive or near-zero gaps, while Asian participants remained below the reference (20.7%). The survey-enriched health status objective produced a substantial reversal in allocation rates across most racial groups. Black (55.9%), Other or Multiple (55.3%), and Hispanic (50.7%) were flagged at higher rates than White participants (44.1%).

**Table 10.**
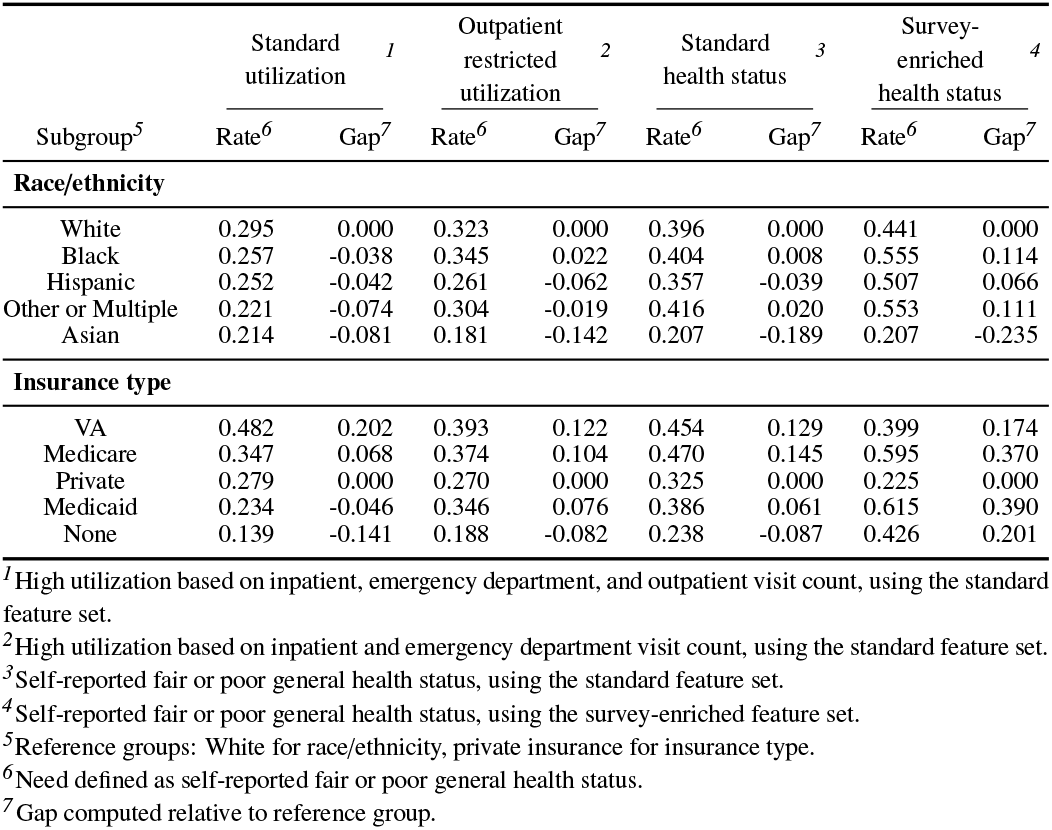
Allocation rate conditional on need and allocation rate gap across training objective conditions, by race/ethnicity and insurance type.

|  | Standard utilization<sup>1</sup> |  | Outpatient restricted utilization<sup>2</sup> |  | Standard health status<sup>3</sup> |  | Survey-enriched health status<sup>4</sup> |  |
|---|---|---|---|---|---|---|---|---|
| Subgroup<sup>5</sup> | Rate<sup>6</sup> | Gap<sup>7</sup> | Rate<sup>6</sup> | Gap<sup>7</sup> | Rate<sup>6</sup> | Gap<sup>7</sup> | Rate<sup>6</sup> | Gap<sup>7</sup> |
| **Race/ethnicity** |  |  |  |  |  |  |  |  |
| White | 0.295 | 0.000 | 0.323 | 0.000 | 0.396 | 0.000 | 0.441 | 0.000 |
| Black | 0.257 | -0.038 | 0.345 | 0.022 | 0.404 | 0.008 | 0.555 | 0.114 |
| Hispanic | 0.252 | -0.042 | 0.261 | -0.062 | 0.357 | -0.039 | 0.507 | 0.066 |
| Other or Multiple | 0.221 | -0.074 | 0.304 | -0.019 | 0.416 | 0.020 | 0.553 | 0.111 |
| Asian | 0.214 | -0.081 | 0.181 | -0.142 | 0.207 | -0.189 | 0.207 | -0.235 |
| **Insurance type** |  |  |  |  |  |  |  |  |
| VA | 0.482 | 0.202 | 0.393 | 0.122 | 0.454 | 0.129 | 0.399 | 0.174 |
| Medicare | 0.347 | 0.068 | 0.374 | 0.104 | 0.470 | 0.145 | 0.595 | 0.370 |
| Private | 0.279 | 0.000 | 0.270 | 0.000 | 0.325 | 0.000 | 0.225 | 0.000 |
| Medicaid | 0.234 | -0.046 | 0.346 | 0.076 | 0.386 | 0.061 | 0.615 | 0.390 |
| None | 0.139 | -0.141 | 0.188 | -0.082 | 0.238 | -0.087 | 0.426 | 0.201 |

<sup>1</sup>High utilization based on inpatient, emergency department, and outpatient visit count, using the standard feature set.
<sup>2</sup>High utilization based on inpatient and emergency department visit count, using the standard feature set.
<sup>3</sup>Self-reported fair or poor general health status, using the standard feature set.
<sup>4</sup>Self-reported fair or poor general health status, using the survey-enriched feature set.
<sup>5</sup>Reference groups: White for race/ethnicity, private insurance for insurance type.
<sup>6</sup>Need defined as self-reported fair or poor general health status.
<sup>7</sup>Gap computed relative to reference group.

By insurance type, uninsured participants showed the most striking allocation rate shifts across objective conditions, moving from the lowest allocation rate of any insurance group under standard utilization (13.9%) to higher rates under outpatient restricted utilization (18.8%), standard health status (23.8%), and survey-enriched health status (42.6%). Medicaid participants showed a similar pattern, with allocation rates increasing across objective conditions (standard utilization: 23.4%, outpatient restricted utilization: 34.6%, standard health status: 38.6%, survey-enriched health status: 61.5%). VA and Medicare maintained positive allocation rate gaps across all objective conditions, with Medicare showing a large positive gap under survey-enriched health status (37.0 percentage points).

## 4. Discussion

### 4.1. Principal findings

This study evaluated how three modifiable algorithmic design choices (selection threshold, feature set, and training objective) shape allocation equity in care management targeting across demographic and socioeconomic subgroups. The findings reveal that equity outcomes are sensitive to design choices in ways that differ by subgroup and condition, and that training objective variation produces the most substantial and consistent equity effects of the three choices evaluated.

The descriptive analysis revealed a systematic misalignment between healthcare utilization and health need across racial, ethnic, and insurance subgroups. Black, Hispanic, and uninsured participants reported fair or poor general health status at higher rates than White and privately insured participants, yet were flagged as high utilizers at substantially lower rates. This pattern is consistent with prior work documenting how structural barriers to healthcare access suppress utilization for disadvantaged populations relative to their health burden, producing a mismatch between utilization-based risk scores and true health need [2, 10, 15].

The baseline equity analysis confirmed that all three models produced consistent FNR and allocation rate disparities across racial, ethnic, and insurance subgroups, with Asian and uninsured participants showing the largest disparities. The Oaxaca-Blinder decomposition further confirmed that the Black-White allocation gap is attributable to differences in observed characteristics between groups, with prior outpatient visits contributing the largest positive endowment and emergency department utilization partially offsetting it. This finding reflects the distinct roles that outpatient and emergency department visits play in healthcare access: outpatient visits are generally discretionary and access-sensitive, subject to suppression by structural barriers such as cost, transportation, and provider availability, while emergency department visits are largely needdriven and less subject to these same barriers [24–27].

Threshold variation produced the most limited equity effects of the three design choices evaluated. Disparities persisted across all threshold conditions for most subgroups, and higher thresholds were associated with widening rather than narrowing gaps, particularly for uninsured and Asian participants. The direction of change in racial EOD also differed by model class, underscoring that the equity implications of threshold policy are likely context- and model-dependent. These findings suggest that expanding program capacity through threshold adjustment is unlikely to serve as a reliable equity intervention in care management targeting.

Feature set variation produced mixed equity effects across subgroups and conditions. Removing all prior utilization variables reduced model discrimination and worsened FNR and allocation rate disparities for Asian and Hispanic participants, while producing modest improvements for Black participants. The outpatient restricted feature set produced a more targeted equity improvement, with Black participants approaching FNR parity under logistic regression, though at higher false negative rates for both groups, a finding directly consistent with the Oaxaca-Blinder decomposition identifying prior outpatient visits as the primary characteristic difference contributing to the Black-White allocation gap. However, this improvement did not extend to Hispanic or Asian participants, suggesting that the outpatient access suppression mechanism operates differently across racial and ethnic groups, and that targeted feature exclusion may produce group-specific rather than universal equity improvements. The survey-enriched feature set produced modest worsening of FNR disparities for Black, Hispanic, and Other or Multiple participants relative to the standard condition, while substantially worsening uninsured allocation rate disparities, suggesting that adding socioeconomic predictors to a utilization-based model does not reliably improve and may worsen allocation equity for the most disadvantaged groups.

Training objective variation produced the most substantial and consistent equity effects of the three design choices evaluated. The outpatient restricted utilization objective, which defined high utilization based on inpatient and emergency department visits only, produced meaningful equity improvements for Black and uninsured participants while maintaining meaningful model discrimination (AUC: 0.820-0.839 across model classes). Switching the training target to self-reported general health status produced more substantial shifts in equity outcomes across subgroups, but at a greater accuracy cost (AUC: 0.688-0.725). The standard health status objective produced the closest clustering of racial EOD values around zero, suggesting that the disparity between utilization-based and needbased targeting reflects a fundamental difference in what the algorithm optimizes for rather than simply a function of predictive accuracy. The pattern identified in the baseline analysis and Oaxaca-Blinder decomposition, in which structural barriers suppress utilization relative to health burden, is partially bypassed when the model is trained to predict self-reported health status directly. The survey-enriched health status objective produced the most pronounced shift, reversing the direction of most racial, ethnic, and insurance-based disparities such that previously disadvantaged groups showed lower FNR and higher allocation rates than their reference groups. Whether this reversal represents an appropriate correction for historical undertreatment or an overcorrection that introduces new forms of inequity is a normative question that depends on the goals of the care management program and the values of the health system deploying it. The substantial accuracy cost associated with the standard health status objective represents a meaningful tradeoff that health systems must weigh against the equity improvements achieved, though the partial recovery of accuracy under the survey-enriched condition suggests that incorporating socioeconomic predictors can help mitigate this cost.

### 4.2 Implications

These findings have several implications for health systems, payers, and algorithm developers seeking to design more equitable care management targeting systems. Most directly, the finding that training objective variation produced the most substantial and consistent equity effects across subgroups suggests that decisions about what an algorithm is trained to predict should be a primary consideration in equitable algorithm design. Where patient-reported health status data are available, leveraging such measures as training targets may offer a meaningful pathway toward more equitable allocation.

The nuanced and subgroup-specific nature of equity effects observed across simulation conditions underscores the importance of systematic evaluation of algorithmic design choices during model development. No single design choice produced uniform equity improvements across all demographic and socioeconomic groups, and several conditions that improved equity for one group simultaneously produced mixed or worsening effects for others. These findings suggest that health systems and algorithm developers should evaluate the equity implications of key structural design decisions across multiple subgroup dimensions simultaneously, as equity assessments focused on a single group or metric may not capture the full distributional consequences of design choices.

These findings also highlight the importance of evaluating algorithmic equity against measures of health need that are independent of the utilization data used to train the model. Utilization-based equity assessments may systematically obscure disparities that arise from structural access barriers, as populations whose utilization is suppressed by those barriers will appear similar to lower-need populations in utilization-based metrics. Access-independent measures of health need, where available, provide a more complete picture of allocation equity and should be incorporated into algorithmic audits of care management targeting systems. Finally, the causal simulation framework developed in this study provides a replicable methodology for evaluating the equity implications of algorithmic design choices during model development, and may be adapted to other healthcare decision contexts and populations in future work.

### 4.3. Limitations

Several limitations of this study warrant consideration. First, all analyses were conducted using data from the NIH All of Us Research Program, which deliberately oversamples historically underrepresented populations and may not be representative of the patient populations served by deployed care management programs. Findings may not generalize to health system or payer populations with different demographic compositions, utilization patterns, or access barrier profiles. Second, the Oaxaca-Blinder decomposition was restricted to the Black-White comparison and was not extended to other racial, ethnic, or insurance-based subgroup comparisons, limiting the scope of the decomposition analysis. Third, the study evaluated three specific design choices and a fixed set of conditions within each simulation, and equity metric point estimates were not accompanied by bootstrap confidence intervals. Other design choices and parameter values not examined here may produce different equity outcomes. Fourth, survey-derived variables, including self-reported general health status used as the access-independent equity benchmark and socioeconomic variables included in the survey-enriched feature set, were collected within the 2017-2019 study period without differentiation between the 2017-2018 baseline period and the 2019 outcome period. Since most All of Us participants complete the survey at enrollment rather than annually, it was not possible to obtain survey responses specific to each study period, potentially introducing temporal misalignment between survey-derived measures and the utilization outcomes they are intended to predict or benchmark. Fifth, estimates for Asian and uninsured participants should be interpreted with caution given the substantially smaller subgroup sample sizes relative to other groups (2.9% and 0.7% of the analytical cohort, respectively), which may contribute to less stable equity metric estimates and limit the interpretability of observed patterns for these groups. Finally, the study examined algorithmic design choices in isolation and did not evaluate interactions between design choices.

### 4.4 Conclusion

This study demonstrates that algorithmic equity in care management targeting is not a fixed property of a deployed model but a consequence of modifiable design choices made during development. Across three systematic simulations, training objective variation produced the strongest and most consistent equity effects, with the self-reported health status objective reducing disparities across most racial and ethnic groups and its survey-enriched variant reversing them. Feature set and threshold variation produced more limited and subgroup-specific effects, underscoring that no single design choice reliably improves equity across all demographic and socioeconomic groups simultaneously. The causal simulation framework developed here provides a replicable methodology for prospectively evaluating the equity implications of algorithmic design decisions, and the access-independent equity benchmark enabled by linkage of electronic health record and survey data offers a more complete picture of allocation equity than utilization-based metrics alone. Together, these contributions offer a foundation for more systematic and equity-conscious algorithm development, where equity is treated as a design consideration rather than a post-deployment correction.

## Acknowledgments

We gratefully acknowledge All of Us participants for their contributions, without whom this research would not have been possible. We also thank the National Institutes of Health’s All of Us Research Program for making available the participant data examined in this study.

## Data availability

This study used de-identified data from the All of Us Research Program’s Registered Tier Dataset (CDRv8 - R9), available to authorized users through the All of Us Researcher Work-bench. Data for this analysis was accessed under an approved Data Use Agreement. As per the program’s Data and Statistics Dissemination Policy, registered users are prohibited from exporting or publicly sharing participant-level information. All data collection protocols for the NIH All of Us Research Program were approved by the NIH All of Us Institutional Review Board (IRB).

## AI Use Statement

During the preparation of this paper, the authors used generative AI (Claude, Anthropic) to assist with the drafting, editing, and refining of the manuscript. The tool had no access to All of Us data and did not generate or compute any study results. The authors reviewed, edited, and take full responsibility for the content of this publication.

